# Estimating the SARS-CoV-2 infected population fraction and the infection-to-fatality ratio: A data-driven case study based on Swedish time series data

**DOI:** 10.1101/2021.05.27.21257900

**Authors:** Andreas Wacker, Anna Jöud, Bo Bernhardsson, Philip Gerlee, Fredrik Gustafsson, Kristian Soltesz

**Affiliations:** Mathematical Physics, Lund University, Sweden; Occupational and environmental medicine, Department of Laboratory Medicine, Lund University, Sweden; Automatic Control, Lund University, Sweden; Mathematical Sciences, Chalmers University of Technology and University of Gothenburg, Sweden; Electrical Engineering, Linköping University, Sweden

**Keywords:** SARS-CoV-2, COVID-19, Sweden, Herd immunity, Healthcare demand prediction, Data-driven modelling

## Abstract

**Aim:** To estimate the COVID-19 infection-to-fatality ratio (IFR), infection-to-case ratio (ICR), and infection-to-ICU admission ratio (IIAR) in Sweden; to suggest methods for time series reconstruction and prediction.

**Methods:** We optimize a set of simple finite impulse response (FIR) models comprising of a scaling factor and time-delay between officially reported cases, ICU admissions and deaths time series using the least squares method. Combined with randomized PCR study results, we utilize this simple model to estimate the total number of infections in Sweden, and the corresponding IFR.

**Results:** The model class provides a good fit between ICU admissions and deaths throughout 2020. Cases fit consistently from July 2020, by when PCR tests had become broadly available. We observe a diminished IFR in late summer as well as a strong decline during 2021, following the launch of a nation-wide vaccination program. The total number of infections during 2020 is estimated to 1.3 million.

**Conclusions:** A FIR model with a delta filter function describes the evolution of epidemiological data in Sweden well. The fact that we found IFR, ICR and IIAR constant over large parts of 2020 is in contrast with claims of healthcare adaptation or mutated virus variants importantly affecting these ratios. The model allows us to retrospectively estimate the COVID-19 epidemiological trajectory, and conclude that Sweden was far from herd immunity by the end of 2020.

## 1 Introduction

The COVID-19 pandemic has posed enormous global challenges to the healthcare sector. To estimate the future need of personnel, equipment and hospital beds, reliable statistical analysis tools are required. Historic data is an important asset in figuring out how to best combine available time series data to gain predictive capability while reducing the influence of bias and other sources of prediction error and uncertainty. At the same time, statistical analysis of the historical epidemic evolution can provide indications for the success of medical treatments and vaccination programs. It also allows estimation of the accumulated number of infections. This number essentially determines the level of herd immunity, and thus received much attention in Sweden during the spring of 2020.

It is a difficult task to predict healthcare, and—of particular interest in the COVID-19 context—ICU demand. This is especially true in an early phase of an epidemic caused by a previously unknown pathogen, such as the SARS-CoV-2 virus that causes COVID-19. While it was possible to falsify several early prediction models based on high sensitivity, *e*.*g*. [1], it remains a largely open question how time series data could be analyzed to arrive at accurate and precise predictions, with practical use to healthcare planners.

We investigate if a particular simple class of time-invariant finite impulse response (FIR) models [2]—those with a delayed delta impulse response—is sufficient to model the relation between time series data. Particularly, our aim is to investigate whether the simple FIR model is sufficient for relating COVID-19 cases (detected infections), ICU admissions, and registered deaths in Sweden. We then demonstrate how such simple models can be used for reconstructing the epidemiological evolution during times of measurement uncertainty caused by limited test capacity, as well as prediction of ICU demand based on case data.

## 2 Methods

### 2.1 Data used

In this paper, the evolution of the pandemic is based on the following official and openly accessible time series reported by the Swedish Public Health Agency:

- *Cases* Daily PCR-confirmed SARS-CoV-2 cases in Sweden. The date refers to the registration.
- *ICU admissions* Daily number of ICU admissions in Sweden for patients with COVID-19 at the given day.
- *Deaths* Daily number of deaths in Sweden for persons with a SARS-CoV-2 infection at the given day.

The data was extracted from [3] on 14 May 2021 and covers dates until 13 May..

Due to delays in reporting, the last 3 weeks for death data and the last 3 days for ICU and case data are disregarded from statistical analysis and displayed by dotted lines in this work. Due to insufficient testing we also omit case data before 18 June 2020 in the model fitting. During the first wave in March–May 2020, PCR-testing was essentially focused to persons admitted to hospital and elderly care in Sweden due to limited testing capacity. In the first half of June, the Swedish government strongly advocated the testing of all persons with symptoms of COVID-19 and supplied financial assistance to the regions as of 11 June 2020. We assume that this had full effect on testing after a further week, which justifies the date given above.

In addition to the *ordinary* testing of persons with suspected COVID-19 infection, results for six *randomized* studies in 2020 and 2021 have been published by the Swedish Public Health Agency [4]. They can be used to estimate the prevalence of COVID-19 in the population at the corresponding times. The studies conducted 24–28 August 2020 and 21–25 September 2020 did not provide any positive samples, while 23, 9, 24, and 43 positive cases where detected for 21–24 April 2020, 25-–28 May 2020, 30 November–4 December 2020, and 12–16 April 2021, respectively. Test results were available for slightly less than 3 000 persons in the studies of 2020 and 4758 persons for the study in 2021. The limited sample size results in statistical uncertainty, indicated by the 68 % confidence limits for the average assuming a Poisson distribution for the number of positively tested. Sampling bias might provide a reduced prevalence for the two latest studies according to the statistical analysis performed in the studies [4]. Here, we use the bare results based on the number of positive cases. For comparison, we also provide an estimate for the ICR based on sampling-bias corrected data.

While deaths and ICU admissions related to COVID-19 naturally also appear in the case data, their total number until mid-May 2021 sums up to only 1.4 % and 0.7 % of the total cases, respectively. Regarding ICU and death data, one has to take into account that approximately 75 % of ICU patients survive [5], and former ICU patients contribute to the death toll with only 13 %, as there are approximately twice as many deaths as ICU admissions. Furthermore, ICU admission data and death data relate to different age groups: While 69 % of the ICU patients are younger than 70 years old, 68 % of the deceased have reached at least the age of 80. (All data from [3] extracted on 14 May 2021.) The small overlap between the groups generating the cases, ICU and deaths time series suggests that statistical correlations between the time series can be expected to reflect the links to their common cause: antecedent SARS-CoV-2 infection in the Swedish society. This motivates the FIR model discussed in Sec. 2.3 with three independent filter functions for cases, ICU admission, and deaths.

Furthermore, data on antibody prevalence from blood donors and health center samples (unrelated to COVID-19-specific testing) have been aggregated and published [6]. Here we provide 95 % confidence intervals for COVID-prevalence based on these data sources.

### 2.2 Parameters used

In addition to the data on the COVID-19 evolution and prevalences provided by the Swedish Public Health Agency addressed in Sec. 2.1, we apply three further parameters within this study:

- The time interval *T*_interval_ = 10 ± 1 days, during which an infected person shows a positive PCR result. See Sec. 2.5 for details.
- The probability *p*_antibody_ = 0.95 ± 0.05, that a previous SARS-CoV-2 infection is detected by an antibody test. See Sec. 3.2 for details.
- The average duration *τ*_*a*_ = 17 between infection and the admission to ICU. See Sec. 3.3 for details.

Note that *T*_interval_ and *p*_antibody_ are used independently of each other in two different determinations of the ICR. Both ways provide essentially the same result, which stabilizes our results against systematic errors in these parameters. *τ*_*a*_ only enters (22) and the time axis in Fig. 3.

### 2.3 Finite impulse response models

Finite impulse response (FIR) models are a class of linear filters. (The interested reader is referred to [2] for a thorough mathematical introduction to FIR and related linear model structures.) They describe the outcome of a time-dependent observable (such as a death rate) by a sum of preceding data (here number of infections at earlier dates), which are weighted by a filter function. They are commonly used for analysing epidemiological problems, where the filter function could represent for example the serial interval distribution. For practical applications, the filter function is often not easy to obtain. Here we show that the assumption of a Dirac delta response, where the filter function has only two free parameters (delay and amplitude), allows for a consistent analysis of the COVID-19 evolution in Sweden.

A central entity for the evolution of an epidemic is the number of new infections 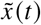, occurring on day *t*. Let 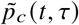, be the probability for an infection starting on day *t* to generate a reported positive PCR test *τ* days later. This results in the observation model

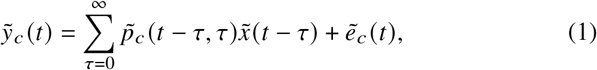

where 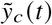 denotes new cases on day *t*, and 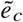 is a zero-mean uncorrelated noise process representing statistical fluctuations associated with the probability 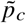. The model has finite impulse since *p*_*c*_ is identically zero for sufficiently large *τ* (*e*.*g*. a human lifetime), and can be regarded practically as zero for *τ* ≫ 1 week. The time index *t* has the unit of days. In (1) it represents that the detection probability distribution, defined through the dependence of the second time index *τ*, may itself vary over time.

The use of the tilde ∼ in (1) is to distinguish unfiltered measurements. Historic observations 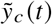 exhibit a clear weekday pattern. For retrospective analysis it is therefore customary to apply a centered 7-day moving average filter,

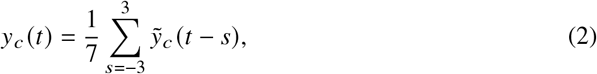

to compensate for such effects. Throughout the paper we will work with time series that have been subjected to filtering according to (2). We will drop the ∼ notation but still write *e*.*g*. “cases” instead of “filtered cases” in favor of readability.

Within linear system theory, a model with the structure of (1) is referred to as a (stochastic) finite impulse response (FIR) model, implying (combined with (2)) that a *y*_*c*_ (*t*) can be described by a finite record of *x* (*t*).

Summation of *p*_*c*_ (*t, τ*) over *τ*, yields the expected infection-to-case ratio (ICR):

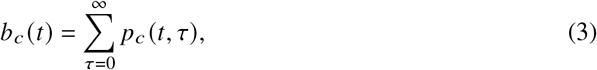

defined as the probability that a person infected on day *t* will eventually become detected and registered as a case.

The central point of the manuscript is that we investigate the hypothesis that *p*_*c*_ (*t, τ*) can be adequately modeled using the delta FIR model

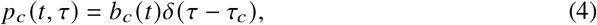

where the discrete delta filter function given by

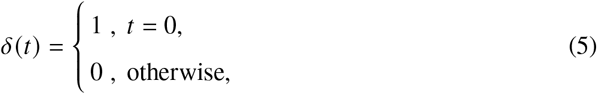

where *τ*_*c*_ is the average delay between infection and case registration. Note that the model (4) is defined for the averaged quantities (2), where *p*_*c*_ (*t, τ*) does not display the weekday fluctuations in *t*, that are likely in 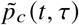. Assuming that the *τ*-dependence of *p*_*c*_(*t, τ*) is reasonably well reproduced by its average *τ*_*c*_ and standard deviation *σ*, the simplified model (4) is justified in App. A. This relies on the assumption that *p*_*c*_ (*t, τ*) does not change on the scale *σ* in *t* and that the second derivative of *x*(*t*) is much smaller than *x* (*t*) / *σ*^2^. For the special case of an exponential evolution for *x* (*t*), this provides an accuracy of better than 5 % if *σ* is less than 46 % of the doubling time, as already stated in [7].

Employing (4), the observation model (1) becomes

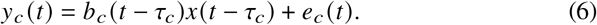

The model (6) asserts that the number of cases *y*_*c*_ (*t*), detected through PCR testing on day *t*, only depends on the number of new infections *x*(*t* −*τ*_*c*_) that occurred *τ*_*c*_ days earlier. Furthermore, the expected dependence is through a linear scaling factor, the ICR.

### 2.4 Relating the time series

We introduce analogous observation models for ICU admissions *y*_*a*_ and deaths *y*_*d*_:

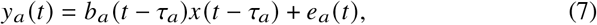

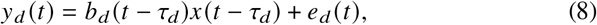

where *b*_*a*_ (*t*) is the Infection ICU admission Ratio (IIAR) and *b*_*d*_ (*t*) the Infection Fatality Ratio (IFR), where the time dependence denotes the infection date.

The underlying infections *x* (*t*) are unknown, and cannot be estimated solely from the measurements *y*_*c*_, *y*_*a*_, *y*_*d*_, since an absolute reference frame against which to estimate the individual time-shifts and gain factors is not available. However, if we disregard the noise terms, we can relate the cases and ICU admissions time series through

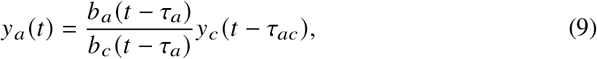

where *τ*_*ac*_ = *τ*_*a*_ − *τ*_*c*_ is the average delay between the registration as a case and the admission to ICU (not necessarily for the same person). Analogously, we have

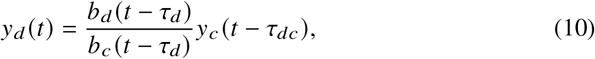

with *τ*_*dc*_ = *τ*_*d*_ −*τ*_*c*_ and

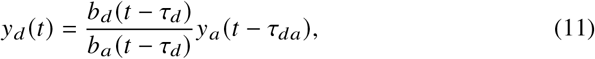

with *τ*_*da*_ = *τ*_*d*_ − *τ*_*a*_.

Eqs. (9–11) can be conveniently fitted to data. If the time-dependence of the *b*-coefficients is negligible, we can fit the ratio *λ* = *b*_*a*_ / *b*_*c*_ and *τ*_*ac*_ from (9) by minimising the sum of squares

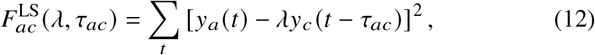

which results in the two fitting parameters *λ, τ*_*ac*_. In order to obtain robust estimates we alternatively minimise the modified sum of squares

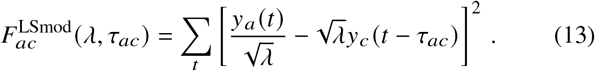

As a third option, we maximize the correlation coefficient

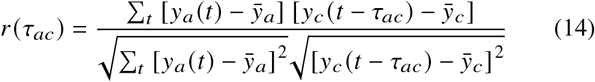

to obtain *τ*_*ac*_ and use 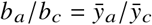. In all three approaches, the times *t* are chosen such that reliable data for both *y*_*c*_(*t* − *τ*_*ac*_)and *y*_*a*_ (*t*) are available. Eqs. (10,11) are treated in the same way, where the indices *a, c* are replaced by *d, c* and *d, a*, respectively, in the formulae above. Note, that each combination of indices applies a different time interval due to the reliability condition.

### 2.5 Calibrating against randomized PCR test data

While (9–11) establish relative relations between the time series *y*_*c*_, *y*_*a*_, *y*_*d*_, a “grounding point” is needed to obtain absolute values of the time shift and scaling parameters of the observation models (6–8). Randomized PCR studies provide such a grounding point, where we use the data discussed in Sec. 2.1. From the number of positive PCR test results in each study, we can estimate the prevalence *N*_positive_ (*t*) by multiplying with the population of Sweden and dividing by the number of tested persons.

The prevalence *N*_positive_ (*t*) depends on the probability *p*_positive_ (*τ*) to have a positive test result *τ* days after becoming infected:

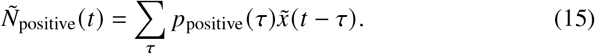

After time-averaging and using again the impulse FIR model this provides

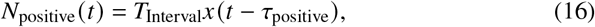

where

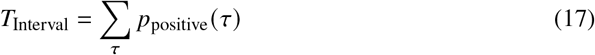

is the average time-interval over which a positive test result is expected and

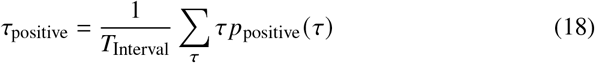

is the average delay after the time of infection. From the data of Fig. 2 of [8], we extract *T*_Interval_ = 10.8 days and *τ*_positive_ = 12 days. Another study [9] found *T*_Interval_ = 9.5 days. Motivated by these numbers we use *T*_Interval_ = 10 ± 1 days and make the simplifying assumption *τ*_positive_ = *τ*_*c*_. The resulting value for *τ*_*c*_ from (22) agrees well with *τ*_positive_ = 12 days, extracted from [8]). Analogously to (9) we fit the relation

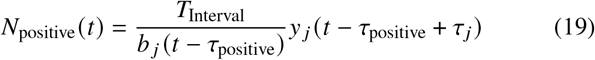

where *j* = *a, d* refers to the data sets for ICU and deaths. We apply all three fitting routines, again neglecting the time-dependence of *b* _*j*_ (*t*) —however, *τ*_positive_ = *τ*_*c*_ is kept fixed. (As we only have 4 non-vanishing data points for *N*_positive_, the use of a second fit-parameter next to *b* _*j*_ could provide spurious results)

## 3 Results

### 3.1 Scaling of data

The symbols on the upper panel of Fig. 1 show the daily Swedish numbers of positively tested persons (named cases), persons admitted to intensive care units (named ICU), and deaths, see Sec. 2.1 for details. These data were averaged over a seven day period (lines) in order to avoid weekday fluctuations, resulting in the functions *y*_*c*_ (*t*) (Cases), *y*_*a*_ (*t*) (ICU admission), and *y*_*d*_ (*t*) (deaths), which are the main data-sets used throughout this article. Here the time *t* is chosen as the central day of the averaging period.

**Fig. 1.**
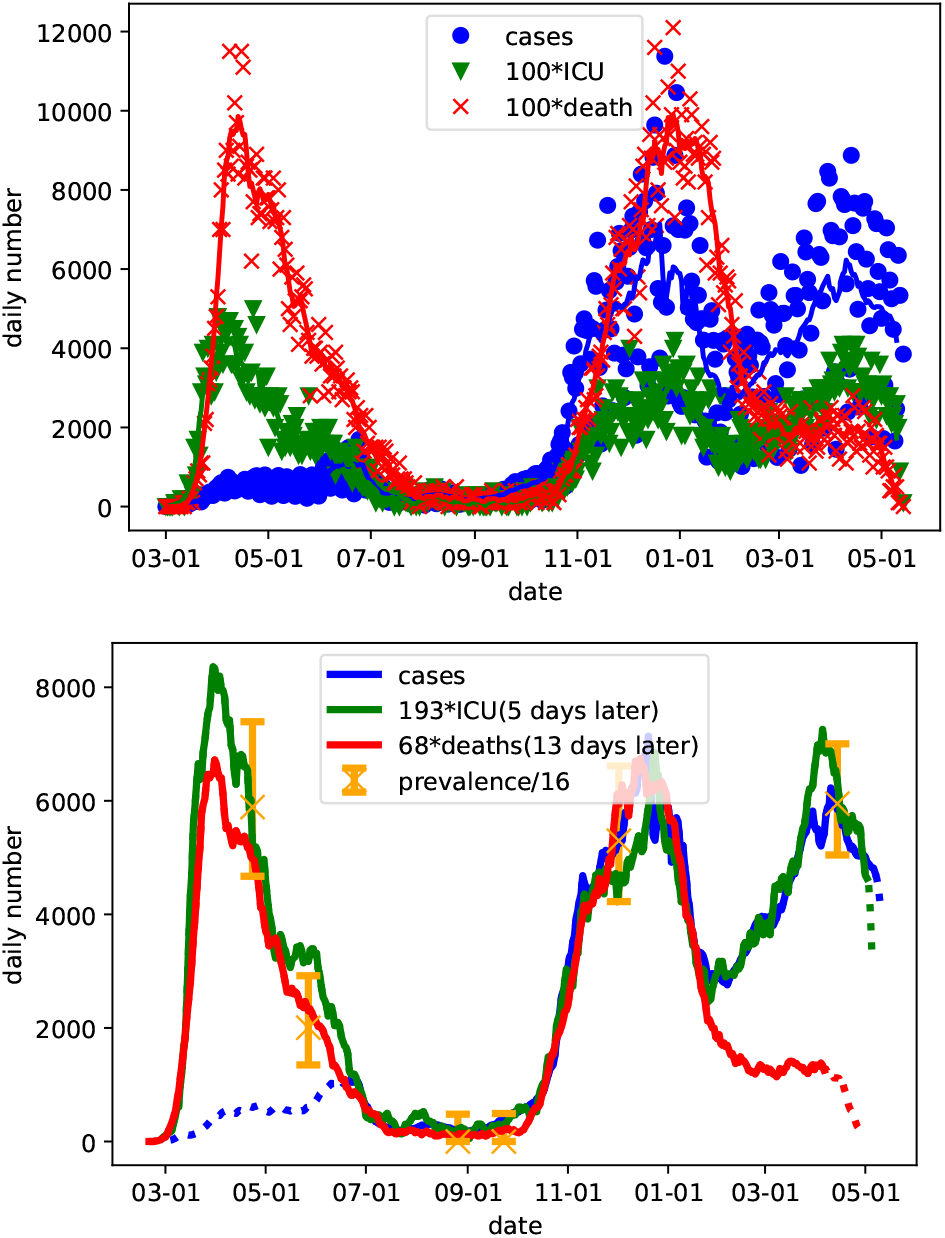
**Upper panel:** Raw data (symbols) used in this study. (The numbers of ICU admissions and deaths have been multiplied by 100 to provide comparable numbers.) The lines show the respective averages over a 7 day period, where the central day of the interval was used in the abscissa. **Lower panel:** 7 day averaged data from upper panel scaled by different factors as given in the legend. The ICU-data are shifted by 5 days to the left and the death-data are shifted by 13 days to the left, so that they essentially fall on one curve. Dotted lines indicate sections of data that are considered as incomplete. The orange error bars provide scaled 68 % (one normal standard deviation) confidence levels of randomized PCR study data. All scaling factors and delays are taken from the LS values in Tab. 1.

As described in Sec. 2.4, we determined the ratios *b*_*a*_ / *b*_*c*_, *b*_*d*_ / *b*_*c*_, and *b*_*d*_ / *b*_*a*_ as well as the corresponding delays by the following procedure. For the fitting, we disregarded the death data after 1 February 2021, as they are affected by the vaccination program and thus a time-dependence for *b*_*d*_ (*t*) is expected here. We also disregard the case data before 18 June 2020, when testing became available to all persons with symptoms in Sweden, see Sec. 2.1. Finally, we consider the data for cases and ICU admission from the last three days and the death data from the last three weeks as unreliable, as late reports are common over these periods. The results are given in Tab. 1. We find that all three approaches provide identical time delays, which we regard as particularly reliable. Also the fractions between the ratios agree fairly well with deviations far below 10 %. In the following we apply the values from the least squares method LS, see Sec. 2.4, but we note, that none of our results depends on this choice. The relations *b*_*a*_ / *b*_*c*_ · *b*_*d*_ / *b*_*a*_ = *b*_*d*_ / *b*_*c*_ and *τ*_*ac*_ + *τ*_*da*_ = *τ*_*dc*_ hold only approximately as different time intervals are used in the fitting due to the exclusion of death data after 1 February 2021 and case data before 18 June 2020 addressed above.

**Table 1.**
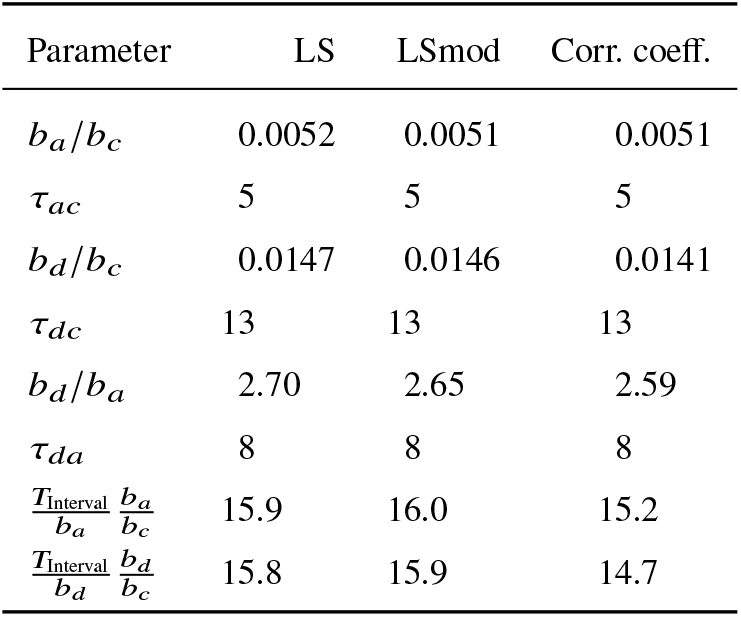
Results for different fitting procedures detailed in Sec. 2.4 for ratios and time delays. In the last two rows the fitted values for *T*_Interval_/*b*_*a*_ and *T*_Interval_/*b*_*d*_ were multiplied with the factors from row 1 and 3 respectively to obtain estimates for *T*_Interval_/*b*_*c*_.

Using these scaling factors *b*_*a*_/*b*_*c*_ = 1/193 and *b*_*d*_/*b*_*c*_ = 1 / 68 as well as the respective time delays, the lower panel of Fig. 1 shows that all three curves agree very well over the second wave of November 2020–January 2021. The ICU and death curve also show a similar behavior at the first wave March–May 2020, albeit the ratio between ICU admission and deaths appears to be slightly higher here. The case numbers are much lower due to the limited testing before mid-July. For the third wave March–May 2021, the ICU and case curve agree very well (the dip in cases around 1 April may be attributed to decreased testing around Easter). We also see that the death curve shows much lower values from around mid-January 2021, which coincides with the start of the vaccination program in Sweden at the end of 2020.

### 3.2 Comparing with randomized PCR and antibody studies

We also fitted the data from the 6 randomized PCR studies to the ICU and death data (here we omitted the sixth study, as the fatality was significantly reduced in 2021, most likely due to the vaccination program) and found almost identical results in both cases, see Tab. 1. Thus, the prevalence of PCR-detectable SARS-CoV2 infections is about 16 times higher than the number of cases (as reconstructed by shifting and scaling the death and ICU data), see the lower panel of Fig. 1. As an infected person can be detected over an average time interval *T*_interval_, but is only registered once as a case with probability *b*_*c*_, this implies *T*_Interval_ / *b*_*c*_ ≈ 15.6 ± 0.9.See Sec. 2.5 for details. Here we used the average of all values provided in the two lowest rows of Tab. 1 and used the maximal deviation as an estimate for the error. The time interval, an infected person is tested positive seems to be less well known. Using *T*_Interval_ = 10 ± 1 days, see Sec. 2.5, this provides the ICR based on the prevalence studies

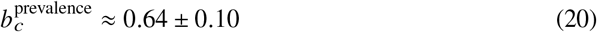

We note that the sampling-bias corrected data for the prevalence results in a slightly larger value 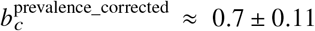 with overlapping error marginal.

The lower panel of Fig. 1 shows, that the case data agree with the prevalences divided by 16 after testing become widely accessible around mid of June 2020. As both the scaled ICU and death data agree well with the prevalences at all times, we can take these curves for estimating the number of cases before mid-July, where the case data are not reliable due to limited testing.

We find a clear plateau of total cases in July–November 2020, as there were few new cases in this period. The average plateau value (at 1. Sept) was 450 000 (reconstructed) cases, with an uncertainty (based on the ICU and death data) of about 55 000. Antibody tests for different groups provide estimates of 700 000 positive persons is Sweden both at the beginning and the end of the plateau, see Fig. 2. This results in *b*_*c*_ / *p*_antibody_ = 0.64 ± 0.08. The probability *p*_antibody_ to develop detectable antibodies has been found to be above 90 % [10], [11]. As it should not exceed 100 %, we assume *p*_antibody_ = 0.95 ± 0.05, and find

**Fig. 2.**
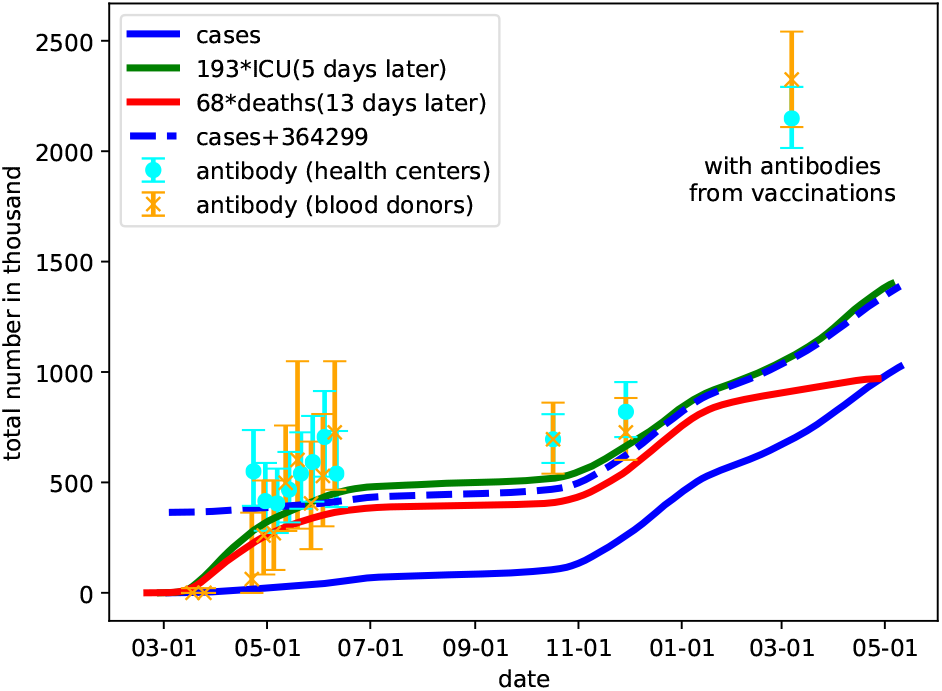
Accumulated number of cases at a given date, where we added the estimates based on scaling the death and ICU data. The dashed blue line provides the case data where we added an estimate for number of cases missed due to limited testing before mid-June. The symbols with error bars show the results of antibody tests performed for blood donors and blood samples from health centers [6], where the fraction of positive tests was multiplied by Sweden’s population. Note that the number of persons with antibodies in March 2021 include detected results from vaccinations.

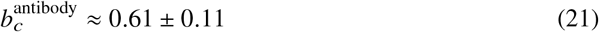

This agrees very well with the different estimate (20), albeit different statistical and systematic errors (in particular the values of *T*_Interval_ and *p*_antibody_) enter both ways to calculate *b*_*c*_. Thus we consider *b*_*c*_ = 0.63 as a good estimate for the ICR with the awareness that a 10 % error is not unlikely.

We note, that a relatively large number of persons with antibodies was found in the study at the beginning of March 2021. Here, one has to take into account that a part of the antibodies detected results from vaccinations. At this time about 700 000 persons had been vaccinated in Sweden and a majority of them should have developed antibodies, when the data was collected.

### 3.3 Reconstructing the IFR and the IIAR

Assuming the ICR *b*_*c*_ = 0.63 for the time after June 18^th^, 2020, we can estimate the variables *b*_*a*_ (*t*) and *b*_*d*_ (*t*) from (9,10). At first we need the absolute delays. Here we rely on the data for ICU admission, which in average occurs about 11 days after the onset of symptoms according to the Swedish Intensive Care Registry [12]. Furthermore, it is known, that it takes about 6 days from the times of infection to develop symptoms [13]–[15]. Thus we use *τ*_*a*_ = 17 days in the following. Based on the values on Tab. 1 we get

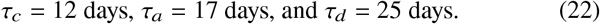

In Fig. 3 we plot the time-dependence of the infection-to-ICU admission ratio (IIAR) *b*_*a*_ (*t*) by a green line on the basis of (9). Here we used 21 day averages to restrict fluctuations. We find that *b*_*a*_ (*t*) ≈ 0.34 % is close to its average, which confirms the quality of the scaling. The larger bump around late July occurs within a range with small numbers of ICU admissions (average of 1.6 in August 2020), so that statistical fluctuations cannot be excluded here.

**Fig. 3.**
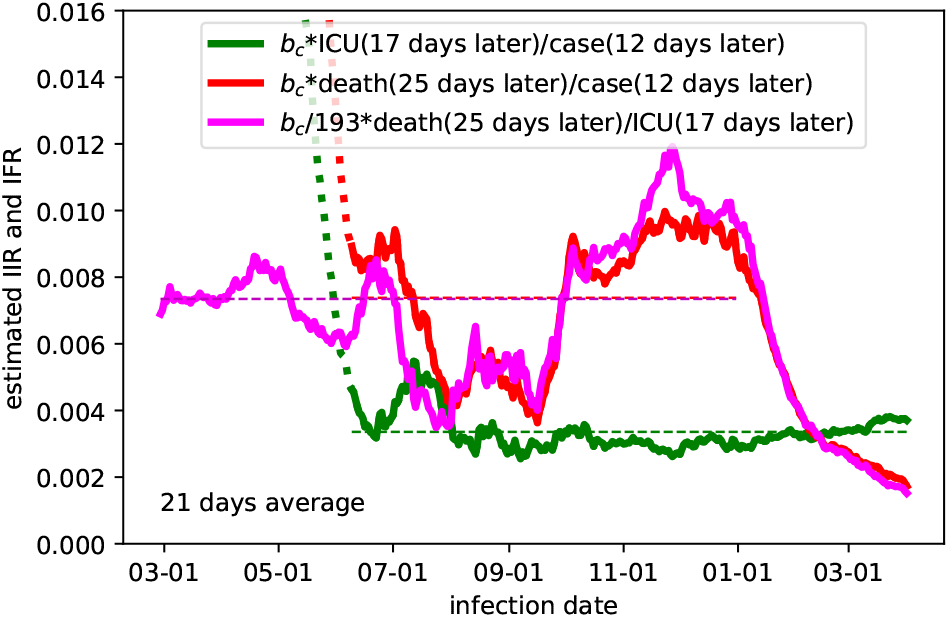
Estimated values for the IIAR *b*_*a*_ (*t*) (green) and the IFR *b*_*d*_ (*t*) (red, magenta) based on (9,10,11). We assume a constant ICR *b*_*c*_ = 0.63, which appears reliable for cases after 18 June (dotted curves indicate that earlier cases data are applied). The horizontal dashed lines provide the average values.

Similarly, we obtain the IFR *b*_*d*_ (*t*) from (10) as shown by the red line in Fig. 3. On average, we have 0.74 % until end of 2020 (before vaccinations started to show effects), but there are pronounced changes over time. For infections in August and September, the IFR is much lower. For infections after 1 January 2021, we see a distinct decline in the IFR reaching values of 0.2 % for infections around 1 April 2021. The same behavior for the IFR can be obtained from from (11) as shown by the magenta line in Fig. 3, which now extrapolates to times before mid-June under the assumption, that the IIAR remained essentially constant in this period as well.

## 4 Discussion

Using openly accessible data from the Swedish Public Health Agency and the Swedish Intensive Care Registry, our approach provides estimates for the infection-to-fatality ratio (IFR), infection-to-case ratio (ICR), and infection-to-ICU admission ratio (IIAR). We find that data for daily cases, daily ICU admission, and daily deaths of individuals with confirmed infection fall on essentially a single curve based on a FIR model with a delta filter function and time-invariant fit parameters, see the lower panel of Fig. 1. There are only two major well-understood exceptions: (I) Cases exhibit a poor match before mid-June 2020, when free PCR testing became broadly available in Sweden for all persons with symptoms. (II) There is a sharp relative decrease in deaths coinciding with the start of the national vaccination program around the turn of the year 2020–2021.

Based on these findings we demonstrate that the approach can be used to retrospectively estimate the cases time series prior to July 2020, that would have been observable with a broad PCR testing program in place. Furthermore, by incorporating data from six randomized PCR studies conducted by the Swedish Public Health Agency and data for the prevalence of antibodies, we obtain an absolute value for the infection-to-case ratio ICR of 0.63 ± 0.07 for the time when testing is easily available to persons with symptoms in Sweden. This value is larger (but compatible within its error) than the value 0.56 obtained in a study for Iceland [10]. It means that approximately 37 % of all infected remained undetected as cases. Regarding systematic errors, the ICR value obtained from our study is reduced if both the time interval for positive testing, *T*_Interval_, and probability to develop measurable antibodies after an infection, *p*_antibody_, turn out to be much less than 10 days and 0.95, respectively. On the other hand, larger values are less likely as *p*_antibody_ = 0.95 was chosen close to its absolute maximum value 1 in the analysis. However, a sampling bias or false-positive tests may allow for deviations, which we cannot judge here.

Fig. 2 indicates that the total number of cases would have been around 820 000 at the end of 2020, if testing in the first half of 2020 had been as available as in the second half. With an ICR of 0.63, this provides about 1.3 million infected persons in Sweden in 2020. This corresponds to 13% of the population and is far below values required to reach herd immunity.

Based on the ICR of 0.63, we obtain the infection to ICU admission ratio IIAR of 0.34% and an average infection to fatality ratio IFR of 0.74 % (before the start of the vaccinations), see Fig. 3. Our value for the IFR is comparable to earlier studies[16]–[18]. Note that possible systematic errors in ICR, as addressed above, affect the IIAR and IFR proportionally. Our analysis shows that the IIAR is rather constant in time, at least for the time after august 2020. In contrast, the IFR varies much stronger over time. We attribute its decline starting for infections around 1 January 2021 to successful vaccination of elderly persons. They dominate the mortality in COVID-19, but are less relevant for ICU admission. However, we lack explanation for the reduced IFR for infections from mid-July to mid-September of 2020.

The close correlation between cases, ICU admissions, and deaths—down to a linear scaling and time-shift once self-test had been made broadly available—warrants further investigation. It could be explained through (I) time-invariance of the ICR, the IIAR, the IFR, (the dip in IFR in summer is not visible in Fig. 1(b) due to the small numbers) and the temporal distributions describing the associated flows; (II) variations in the mentioned entities that have essentially cancelled each other throughout 2020; (III) external confounders providing this canceling effect. Case (I) would imply that the healthcare system’s ability to save COVID-19 patients and the impact from different virus mutations has not changed markedly. Case (II) would be surprising for the time from August 2020, where the ratio between cases and ICU admission is largely constant in time, see Fig. 3. Thus it is most likely that *b*_*c*_ (*t*) and *b*_*a*_ (*t*) are both constant in this range. However, for the first half of 2020, things are less obvious. The variations observed in the magenta curve may result from reductions in *b*_*a*_(*t*) and *b*_*d*_(*t*) (*i*.*e*. improvement in healthcare) occurring at different times. Case (III) would also be noteworthy since the groups of deceased and persons admitted to ICU care have marginal overlap with each other.

The studied time series alone are not sufficient to map out the causation of the observed correlations, thus distinguishing between (I) and combinations of (II) and (III). However, an understanding of the underlying mechanism could be obtained by retrospectively tracking individual traces connecting the considered time series (*e*.*g*. persons who have tested positive, been admitted to an ICU or died with COVID-19). Importantly, if the correlation can be understood and shown not to be coincidental, it could constitute the basis for an accurate 1–2 week predictor of the ICU demand.

## 5 Conclusion

The summarizing conclusion from our observations is that important insight into numerous aspects of an ongoing epidemic can be obtained by considering the scaling between different time series, where time shifts are crucial. This is based on an FIR model with a delta filter function, which is shown to work well. We demonstrated this by reconstructing the daily number of cases for the first half year of 2020 periods, where testing was limited in Sweden, and extracting time variations in the infection fatality ratio.

## Data Availability

Only publicly available data sets have been used. They are referenced in the manuscript.

## Acknowledgements

This work was partially funded by the ELLIIT Strategic Research Area.

## Conflict of interest

The authors declare no conflict of interest.

## A Quality of the delta-impulse model

After averaging to remove weekly fluctuations and neglecting fluctuations *e*_*c*_ (*t*), (1) provides

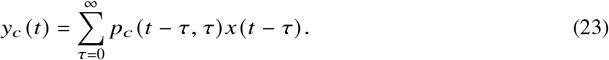

While we do not know much about *p*_*c*_ (*t, τ*), we make the reasonable assumption that it has a single peak and does decay rather quick for large delays *τ*. In this case it is reasonably represented by its average 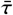 and standard deviation *σ* defined as

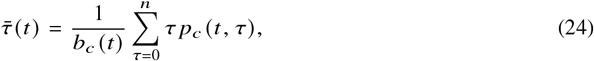

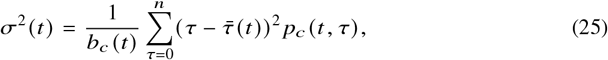

where we used the normalisation from (3). Assuming, that *x* (*t*) is a smooth function, which appears reasonable after 7-days averaging, we may approximate by a second order Taylor expansion

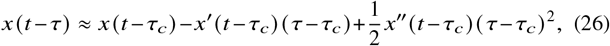

where, for given *t*, the average delay *τ*_*c*_ satisfies 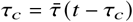 Then we obtain from (23) that

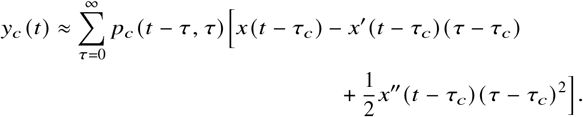

Assuming *p*_*c*_ (*t* − *τ, τ*) ≈ *p*_*C*_ (*t* − *τ*_*C*_, *τ*) as the detection probability should not change within a time-scale of *σ* for constant delay *τ*, we find

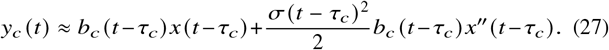

The first term is just our delta-impulse model, see (4), while the second term provides a correction less than 5 % if

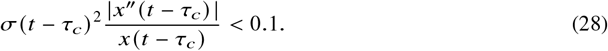

If the infections show an exponential behavior *x*(*t*) = *x*_0_*e*^*rt*^, this provides *σ r <* 0.316 or *σ <* 0.46*t*_doubling_ with the doubling time *t*_doubling_ = ln 2/*r*. The last relation was stated in Ref. [7] for the applicability of the delta-response for exponential evolutions. Here we generalised this to arbitrary evolutions *x*(*t*), where *t*_doubling_ / ln 2 is replaced by the square root of the inverse relative second derivative.

## Nomenclature

**Acronyms**
FIR: Finite impulse response [model]
ICR: Infection-to-case ratio
ICU: Intensive care unit
IFR: Infection-to-fatality ratio
IIAR: Infection-to-ICU admission ratio
LS: Least-squares [method]
PCR: Polymerace chain reaction

## Subscripts and modifiers

*a*: Infection-to-ICU admission
*c*: Case (detected infection)
*d*: Infection-to-fatality (death)
∼: Weekly central moving average

## Variables and symbols

*δ*: Dirac’s delta distribution
*γ*: IIAR/ICR
*σ*: Standard deviation
*τ*: Time delay (unit:days)
*b*: Gain parameter
*e*: Error or residual time series
*N*: Number of individuals
*p*: Probability
*t*: Time index (unit:days)
*x*: New infection time series (not directly observable)
*y*: Daily time series (observation)

## Notes

### Competing Interest Statement

The authors have declared no competing interest.

